# Healthcare Providers’ Satisfaction and associated factors with clinical laboratory service at government hospitals in Southwest Shewa Zone Oromia, Ethiopia

**DOI:** 10.1101/2023.07.25.23293175

**Authors:** Sintayehu Asaye, Getachew Alemu, Dejene Gebre, Lule Teshager, Girma Yadesa

**Affiliations:** Jimma University, School of Medical Laboratory Sciences, Jimma,Ethiopia; Tulu Bolo General Hospital, South west, Ethiopia

**Keywords:** Healthcare providers, Satisfaction, Government Hospital, Southwest

## Abstract

**Background:** Healthcare providers are vital clients of the clinical laboratory. Problems in laboratory services have a significant impact on healthcare providers’ diagnosis, intervention, or prevention strategies during patient healthcare. Healthcare providers’ satisfaction wasn’t assessed in the study area. So, the study aimed to evaluate their satisfaction, which helps improve laboratory services and serves as a prerequisite for accreditation.

The study aims to assess healthcare providers’ satisfaction associated factors with laboratory services among all government hospitals in the southwest Shewa zone, Oromia, Central Ethiopia 2022.

**Methods:** A cross-sectional study was conducted on 314 healthcare providers from June to July 2022. Healthcare providers who met the eligibility criteria and volunteers were included. Before being exported to SPSS version 25, the data were coded and entered into Epi Data version 3.1 for completeness checks. After descriptive statistics were performed, variables were subjected to bivariate and multivariable logistic regression analysis to identify associated variables, and variables with a p. value < 0.05 were considered statistically significant. Finally, the results were shown in the text and table.

**Results:** According to the study, 54 % of healthcare providers were satisfied with laboratory services. The assistance of the lab. Handbook (AOR=1.676, 95% CI=1.002, 2.801), notification of the change in services (AOR=1.735, 95% CI=1.029, 2.925), getting urgent test results in time (AOR=2.349, 95% CI=1.39, 3.68), courtesy of lab. Staff (AOR=1.924, 95% CI=1.115, 3.321), reliable/quality of lab. result (AOR=3.69, 95% CI= 2.083, 6.538), and consistency of laboratory quality services (AOR=1.706, 95% CI=1.012, 2.875) were factors significantly related to satisfaction.

**Conclusion:** About half of the healthcare providers were dissatisfied with laboratory services. Therefore, to increase the satisfaction of healthcare providers’ hospital administrative and laboratory staff, they should work together on the identified factors. In order to determine overall clients’ satisfaction with laboratory services, an assessment of patient satisfaction will be required

## Background

Healthcare providers’ satisfaction refers to the state of pleasure or happiness of healthcare providers with the laboratory services that were previously desired for improved clinical management of patients. Measuring healthcare providers’ satisfaction is an integral part of a laboratory quality management system; furthermore, it encompasses patient management up to hospital administration strategies across the globe (1-3).

Clinical laboratories have distinctive sorts of clients. Clients of medical laboratories can include healthcare providers, patients, public health organizations, and the community (4-5). Healthcare providers are considered the vital or central clients of the clinical laboratory. Healthcare providers want laboratory test results that are reliable, accurate, and relevant for patient care (6).

Assessing healthcare providers’ satisfaction with clinical laboratory services benefits the laboratory and is required for global clinical laboratory measurements, which may be a prerequisite for laboratory accreditation. Healthcare providers arrange tests, collect samples and make clinical decisions based on test results (7-9).

Physicians and nurses are highly dissatisfied with laboratory services, as shown in different studies. Sixty percent of physicians were dissatisfied with laboratory services, according to a study done by the College of American Pathologists’ Q-Probes Study of 81 institutions. On the other hand, the study done on client satisfaction with clinical laboratory phlebotomy services at a Tertiary Care Unit level, conducted in South Korea in 2012, revealed that 49.1% of customers were dissatisfied (10,11).

According to a national survey conducted in Ethiopia at public hospitals, almost half of the physicians were dissatisfied with laboratory service (8). Also, the study conducted in western Ethiopia shows that 49.3% of healthcare providers were dissatisfied (12).

This study will provide information on overall healthcare providers’ satisfaction with laboratory services at government hospitals in the Southwest Shewa Zone, central Ethiopia. Furthermore, the findings will assist the study hospitals in identifying gaps and achieving better laboratory service.

## Methods

### Study area and period

The study of healthcare providers’ satisfaction with clinical laboratory services was conducted at government hospitals in the southwest Shewa zone, Oromia, central Ethiopia from June to July 2022. Southwest Shewa Zone has a population of 1,101,129 people according to the 2007 census conducted and is located 118 kilometers southwest of Addis Abeba. Southwest Shewa is bounded by the Addis Ababa Special Zone and East Shewa Zone in the north and northeast, the Gurage Zone of the South Nation Nationality and Peoples Region in the south and west, and the West Shewa Zone in the North and West (13). The zone had five government hospitals with five central laboratories.

### Study Design

Institutional-based cross-sectional study design was conducted.

### Source population

All health worker at government hospitals in the Southwest Shewa Zone, Oromia, central Ethiopia.

### Study population

Healthcare providers who voluntarily participated and who fulfilled the inclusion criteria.

### Inclusion criteria

Healthcare providers who utilize clinical laboratory service results to improve clinical management for their patients.

### Exclusion criteria

- Healthcare providers with work experience under six months.
- Healthcare providers with working experience longer than six months but less than three months at study hospitals.
- Healthcare providers on maternity leave were excluded.

### Sample size determination

To have an adequate sample size all healthcare providers who have been working at Tulu Bolo General Hospital (102), Waliso General Hospital (67), Bantu Primary Hospital (47), Lemen Primary Hospital (49), Hamaya Primary Hospital (49), were included. Identification of the potential candidates using the inclusion criteria was done first before data collection to attain the final individuals.

### Data collection instruments

The self-administered, structured questionnaires consisted of five socio-demographic characteristics and twenty-seven levels of satisfaction variables. A total of 21 levels of satisfaction variables were categorized into four service quality dimensions (tangibility, responsiveness, assurance, and reliability), and six levels of satisfaction variables were categorized as the overall satisfaction component. Based on the number of healthcare providers available in each hospital, the self-administered structured questionnaires were distributed to five hospitals. After obtaining oral consent from healthcare providers, the self-administered, structured questionnaires were distributed to them. If there was any confusion on the questionnaire during data collection, the data collector, supervisor, and principal investigator were tasked with explaining the situation to the study participants. Moreover, the principal investigator supervises the overall process of data collection. Finally, the complete questionnaire was collected at the end of each day.

#### Data quality management

Nurses with a bachelor’s degree collected the data. Prior to actual data collection, the principal investigator provided a day of training to ten data collectors and five supervisors on data collection procedures, the purpose of the study, the contents of the questionnaire, and ethical issues concerning study participants. Before the start of the study, the principal investigator ensured that the data collectors were competent. This was accomplished by checking the understanding of data collectors about each item and assessing the accuracy of filling out questionnaires. The supervisor checked the method of data collection and the completeness of the answers to the questions. The principal investigators closely followed up on the process of data collection and were given an immediate solution if there was any confusion on the questionnaire.

Self-administered structured question was pre-tested before the actual data collection at Guder General Hospital was selected for the pre-test because it had similar characteristics to the study area. Five percent of healthcare providers (18) participated in the pre-test this was to check whether the question were simple, clear, and easily understandable. Based on the comment and feedback of pre-testing the questionnaire was revised.

#### Data processing and analysis

Data were exported to the Statistical Package for Social Sciences (SPSS) version 25 software after the completeness of the collected data was proven, coded, and entered into Epidata version 3.1. Simple descriptive statistics, such as frequency and percentages were used to assess socio-demographic and level of satisfaction variables for healthcare providers’ satisfaction with clinical laboratory services.

After the binary logistic regression assumption was fulfilled, logistic regression analysis was implemented. A bivariate logistic regression was initially carried out to determine the associated factors for each independent variable with the outcome variable. To adjust for confounders, all variables significant with p. value < 0.25 in the bivariate analysis were shifted to the multivariable logistic regression model.

Variables adjusted Odds Ratio (AOR) and p. value < 0.05 at 95% confidence interval during multivariable logistic regression analysis was considered as statistically significant with the satisfaction of healthcare providers. Adjusted Odds Ratio (AOR) with a 95% confidence interval (CI) was used to quantify the strength of the association between associated factors and healthcare providers’ satisfaction. The Hosmer and Lemeshow goodness of fit test was used to determine the final model’s adequacy, which was (p = .095). The assumption of multi-colinearity was checked (VIF < 5). In addition, the backward method and first reference categories were applied in multivariable logistic regression for the selection of statistically significant variables. Finally, quantitative results were presented using descriptive statistics in the texts and table. Likewise, qualitative data that was gathered through, observation checklists from the laboratory were summarized according to their particular thematic area.

## Results

A total of 314 healthcare providers participated in the study, with a response rate of 92.4%. The majority were nurses (63.7%), and around 32.5% of the participants were from Tulu Bolo General Hospital. Other participants were: 21.3% from Waliso General Hospital, 15.6% from Hamaya Primary Hospital, 15.6% from Leman Primary Hospital, and 15% from Bantu Primary Hospital.

The age of 46.8% of healthcare providers was in the range of 20–29 years, of which 60.2% were men. Nurses made up 63.7% of all study participants, with midwives accounting for 16.2%. The participants of the study served for a minimum of six months, with the majority (56.0%) serving for more than five years.

The highest satisfaction of healthcare providers with laboratory services was from the emergency patient department while the lowest satisfaction was from the inpatient department.

Overall satisfaction of healthcare providers with clinical laboratory services was 54%, with a 95% confidence interval of 48.5-59.7.

### Factors affecting satisfaction of healthcare providers with laboratory services

The satisfaction and quality of laboratory test results were found to be significantly related. The quality/reliability of laboratory test results will boost satisfaction by 3.69 times having (AOR = 3.69, 95% CI (2.083-6.538), P. value < .001). Obtaining urgent laboratory test results will 2.349 times increase satisfaction having (AOR = 2.349, 95% CI (1.39–3.68), P. value =.001). Laboratory staff courtesy is significantly related to satisfaction, will increase satisfaction by 1.924 times having (AOR = 1.924, 95% CI (1.115-3.321), P value =.019). (**Table 1**)

**Table 1:** Predictor variables of the healthcare providers’ satisfaction toward clinical laboratory services at the Government Hospital in southwest Shewa zone, central Ethiopia July 2022. (N=314)

| Variables | Category | Healthcare providers' |  | COR<br>(95% CI) | P. value | AOR<br>(95%CI) | P. value |
| --- | --- | --- | --- | --- | --- | --- | --- |
|  |  | Sat N (%) | Dis N (%) |  |  |  |  |
| Assistance of lab. Handbook | No | 67(21.3) | 80(25.5) | 1 |  | 1 |  |
|  | Yes | 103(32.8) | 64(20.4) | 1.9(1.2,3) | .004 | 1.7<br>(1,2.8) | .049 |
| Courtesy of laboratory staff | No | 41(13.1) | 64(20.4) | 1 |  | 1 |  |
|  | Yes | 129(41.1) | 80(25.5) | 2.5(1.5,4.1) | .001 | 1.9<br>(1.1,3.3) | .019 |
| Notification changes in services | No | 55(17.5) | 78(24.8) | 1 |  | 1 |  |
|  | Yes | 115(36.6) | 66(21) | 2.5(1.6,3.9) | <.001 | 1.7(1,2.9) | .039 |
| Getting urgent test result in time | No | 469(14.6) | 80(25.5) | 1 |  | 1 |  |
|  | Yes | 124(39.5) | 64(20.4) | 3.4(2.1,5.4) | <.001 | 2.3(1.4,3.7) | .001 |
| Quality/ reliability of test result | No | 27(8.6) | 73(23.2) | 1 |  | 1 |  |
|  | Yes | 143(45.5) | 71(22.6) | 5.4(3.2,9.2) | <.001 | 3.7(2.1,6.5) | <.001 |
| Consistent of laboratory quality services | No | 64(20.4) | 81(25.8) | 1 |  | 1 |  |
|  | Yes | 106(33.8) | 63(20.1) | 2.1(1.3,3.3) | .001 | 1.7(1, 2.8) | .045 |

**Abbreviations:** Sat= satisfaction; Dis= dissatisfaction; COR= crude odd ratio; CI= confidence interval; AOR=adjusted odd ratio, N= number of healthcare providers.

Notification of changes in services will increase satisfaction by 1.735 times having (AOR = 1.735, 95% CI (1.029-2.922), p.value .039). Quality service consistent will increase satisfaction by 1.706 times having (AOR = 1.706, 95% CI (1.012-2.875), P. value =.045). Assistance of laboratory handbook for utilization of laboratory services will 1.676 times increase satisfaction having (AOR = 1.676, 95% CI (1.002–2.801), P. value =.049). (**Table 1**)

## Discussion

Overall, the objective of this study was to assess the level of healthcare providers’ satisfaction and associated factors with clinical laboratory services among government hospitals in the Southwest Shewa Zone, Oromia Region, Central Ethiopia. The study covered all government hospitals in the Southwest Shewa Zone using twenty-seven levels of satisfaction variables that cover various aspects of laboratory services.

The study revealed that the overall satisfaction level of healthcare providers was 54%, which was consistent with the study conducted in Northwest Ethiopia at 51.1% (14), and West Ethiopia at 50.7% (12), but inconsistent with studies conducted in Nekemte, Ethiopia, at 62.8% (15), and Saudi Arabia at 64.9% (16). This lower result of current study might be due to the current study’s was large sample size, the low human power of laboratory personnel at the study hospitals the current study’s used large data collection tool, and moreover, the fact that the current study was conducted at three primary and two general hospitals, whereas the Nekemte Ethiopia and Saudi Arabia studies were conducted at a single referral hospital.

According to this study, 59.9% of healthcare providers were satisfied with getting urgent test results. Furthermore, the study’s findings revealed that getting an urgent test result is significantly related to satisfaction and will increase the satisfaction of healthcare providers by 2.3 times. This finding is relatively consistent with the previous studies done in south Ethiopia (57.4 %) (17), and west Ethiopia (55.6%) (12). but inconsistent with the study done in Saudi Arabia (67.5%) (16). The discrepancy can be due to the lower number of laboratory employees and workload at current study hospitals, as well as the type and number of study facilities involved in the current study.

The results of this study reveal that 45.9% of healthcare providers were satisfied with the routine test turnaround time of laboratory services. This finding is more satisfactory when compared with the previous study conducted in Yemen and Ethiopia. Routine TAT tests dissatisfied 67.1% of customers in Yemen (18) and 60.4 percent of customers in West Ethiopia (12). According to the study, routine tests (TAT) were the most common cause of dissatisfaction with clinical laboratory services. This highest dissatisfaction with the result was due to a lack of monitoring TAT at current study hospitals, low human power in the study facilities (**Table 1**), and a high flow of patients, so many patients’ test results were put together and reported at once. A solution that can reduce TAT for both emergency and routine TATs is to start testing services in inpatient and emergency departments and at point-of-care testing, which is performed by a nurse.

## Conclusions

The results of the study indicate that about half of healthcare providers were satisfied with the laboratory services. Additionally, the study identifies that the assistance of the laboratory handbook for laboratory service utilization, the courtesy of laboratory personnel, notification of changes in services, getting urgent test results on time, quality/ reliability of results, and consistency of quality services were factors that affect the level of satisfaction of healthcare providers. Therefore, in order to address the issue raised in the study, hospital administration, healthcare workers, and laboratory personnel should work more closely together and achieve better laboratory services.

## Data Availability

Data will be available upon the convincing request from the corresponding author.

## Declaration

### Ethics approval and consent to participate

Ethical approval was received from Jimma University’s Institute of Health Ethical Review Board before data collection. Following approval, the School of Medical Laboratory Science wrote a letter of support to the southwest Shewa zonal health office and studies hospital. To end, the study’s objective was briefly explained, and an agreement was made with the study site that the data would be kept private and utilized only for research purposes. Each participant was notified about the objective of the study, the right to refuse or participate in the study, and the secrecy and confidentiality of the information. They were assured that they wouldn’t be penalized for not participating and that their responses to the questions would not affect them. Eventually written informed consent was taken from all study participant voluntarily just before the start of data collection.

### Consent for publication

not applicable

### Availability of data and Materials

Data will be available upon the convincing request from the corresponding author.

### Authors’ contributions

All authors have made a significant contribution to the conception, study design, acquisition, data analysis and interpretation of the results. They also participated in drafting the manuscript, critically reviewed and had consensus on the journal to which the article was to be submitted. All authors read and approved the final version of the manuscript and agreed to be accountable for all the contents of the manuscript.

## Acknowledgments

We would also like to thank data collectors, study participants, and supervisors as well as those who directly or indirectly contributed to this study.

